# Impact of *AKR1C2* and *AKR1C3* single nucleotide polymorphism rs28571848 in adipose tissues of individuals with severe obesity

**DOI:** 10.1101/2025.10.10.25337731

**Authors:** Giada Ostinelli, Alan Ramalho, Marie-Fréderique Gauthier, François Julien, Laurent Biertho, Marie-Claude Vohl, André Tchernof

## Abstract

**Background:** Adipose tissue androgen turnover, dictated at least in part by the enzymes AKR1C2 and AKR1C3, has been linked to abdominal obesity. Recently, we investigated a single-nucleotide polymorphism (SNP) named rs28571858, that might increase *AKR1C2* and *AKR1C3* expression in human adipose tissue. Here, we studied the impact of rs28571848 on adipose tissue function and cardiometabolic health in bariatric surgery candidates.

**Methods:** We genotyped a sample of 2776 bariatric surgery candidates and retrospectively obtained anthropometry, blood lipid and glucose profiles, menopausal status and medication use. In a subsample of 135 individuals (62% women, age 42 years, BMI of 51 kg/m^2^), we additionally assessed *AKR1C2* and *AKR1C3* expression in whole tissue by RT-qPCR. Features of adipose tissue dysfunction, such as mean adipocyte diameter and pericellular fibrosis were assessed by histological staining and semi-automated image analysis. Finally, adipose tissue AKR1C family enzyme activity was measured by fluorimetry.

**Results:** The rs28571848 SNP affected *AKR1C3* expression in both subcutaneous (SAT) and visceral adipose tissue (VAT) in women only, while not altering *AKR1C2* expression in either men or women. Individuals carrying the minor allele exhibited increased VAT AKR1C activity compared to those with the wildtype genotype. Analysis of blood lipid profile in the whole cohort revealed that “TT” carriers had elevated total cholesterol, LDL-cholesterol, and indices of insulin resistance.

**Conclusion:** The rs28571858 SNP increased adipose tissue *AKR1C3* expression and activity in women. Increased AKR1C3 may contribute to an adipose tissue milieu that prompts lipogenesis, adversely affecting cardiometabolic health by disrupting lipid homeostasis and insulin sensitivity.

**NEW AND NOTEWORTHY:**

- rs28571858 has a Minor Allele Frequency (MAF) of 18.7% in a cohort of 2776 residents of Quebec, Canada.
- rs28571858 is associated with adipose tissue *AKR1C3* expression and activity in women.
- Minor allele homozygotes carriers have increased total cholesterol, LDL-cholesterol, and indices of insulin resistance.

## INTRODUCTION

A large number of studies demonstrated how body fat distribution outweighs body mass index (BMI) as a predictor of the cardiometabolic complications associated with obesity (1). Indeed, excessive visceral adipose tissue (VAT), located in the visceral compartment, i.e. around the mesentery and the greater omentum, is directly linked to increased cardiometabolic risk factors (2). Population studies illustrate how excessive VAT increases the odds ratio of cardiovascular diseases (3) to a greater extent than epicardial fat (4). Similarly, excess visceral fat rather than total adiposity, is an independent predictor of type 2 diabetes mellitus (T2D) incidence (5). Recent mendelian-randomization studies additionally identified body fat distribution, particularly visceral adiposity, as the main driver of insulin resistance and thus T2D (6). The same group further demonstrated that waist circumference (WC), a marker of VAT excess, was strongly associated with fatty liver disease, T2D and coronary artery disease (7).

Sex hormones have long been studied as possible drivers of the body fat distribution differences observed between men and women. An extensive review of the literature reported that in men, hypoandrogenemia is associated with increased visceral adiposity, while the opposite is true for women (8). Studies using genome-wide association (GWAS) or mendelian randomization further support the role of circulating testosterone levels in adiposity and T2D but additionally stressed the importance of sex stratification (9, 10).

In adipose tissue, androgens such as testosterone and dihydrotestosterone, can either enter the tissue via the circulation or be synthetized *in situ* (11). However, we (12) and others (13), have demonstrated that the contribution of plasma to the adipose tissue pool of androgens is minimal. This stresses the importance of adipose tissue steroidogenic enzymes in the local regulation of the androgen pool. The conversion of androstenedione to testosterone is under control of 17β-hydroxysteroid dehydrogenase type 5 (17β HSD5, also known as aldo keto reductase 1C3, AKR1C3), while 5α-reductase is responsible for the reduction of testosterone into dihydrotestosterone (DHT) (11). On the other hand, androgen deactivation and more precisely DHT reduction is under the control of 3α-HSD type 3, also known as AKR1C2 (14). *AKR1C2* and *AKR1C3* are both highly expressed in adipose tissue (15), and their expression in preadipocytes increases with differentiation (16). We and others have demonstrated that these two enzymes are associated with obesity, and in particular with visceral adiposity (17–19). Our group has repeatedly shown that *AKR1C2* expression and activity are higher in women with excess VAT (20), which was further supported by the association between *AKR1C2* and adipocyte diameter (12, 21).

Recently, we identified a region on chromosome 10 containing several single nucleotide polymorphisms (SNPs) that can alter the expression of *AKR1C2* and *AKR1C3* (19). Due to high linkage disequilibrium in this region, we were unable to isolate a single possible causal SNP; however, we identified rs28571848 as a strong candidate (19). Here, we investigate 135 bariatric surgery candidates with the rs28571848 SNP, which has been suggested to increase the expression of *AKR1C2* and *AKR1C3* in human adipose tissue. Our aim was to establish the impact of rs28571848 on markers of adipose tissue function and cardiometabolic health.

## METHODS

### Participants

Individuals with severe obesity (BMI ≥ 35 kg/m^2^), being at least 18 years old, and being eligible for elective bariatric surgery were recruited to be included in the Quebec Heart and Lung Institute Biobank. Eligibility for this study was considered if the patient consented to sample donation and genetic testing. *A priori* power analyses, based on previous data (12), were made using G*Power, and indicated that a sample of 38 individuals per genotype group was required to identify a difference in VAT AKR1C2 activity and visceral adiposity. In addition, based on publicly available data, we predicted a minor allele frequency (MAF) to be around 15% (19), allowing the identification of 68 “TT” carriers in a sample of approximately 3000 participants.

Blood samples of 2798 participants were obtained from the Quebec Heart and Lung Institute Biobank. Following genomic DNA extraction from the blood buffy coat (GenElute Blood Genomic DNA kit, Sigma), we used TaqMan® SNP Genotyping Assays (Thermofisher) to discriminate between rs28571848 carriers. After the exclusion of invalid reads (N=15) and individuals from which medical records were no longer available (N=7), we obtained a sample of 2776 participants, among which 99 were “TT” carriers. Participants with available adipose tissue (SAT and VAT) at the Quebec heart and Lung Institute Biobank were matched for age, sex, BMI and date of surgery. This allowed us to obtain a sample of 45 participants, including 17 men and 28 women in each genotype, labelled “CC”, “CT” and “TT” (total sample N=135).

Participant characteristics can be found in **Table 1**. Blood profiles (including lipids, glucose-insulin homeostasis), medication use, menopausal status, anthropometry, liver histology and type 2 diabetes (T2D) as well as liver steatosis diagnosis were retrieved from medical records. Homeostatic Model Assessment of Insulin Resistance (HOMA-IR) was calculated as follows: Fasting Glucose (mM) * Fasting Insulin (mU/L) / 22.5.

**Table 1:**
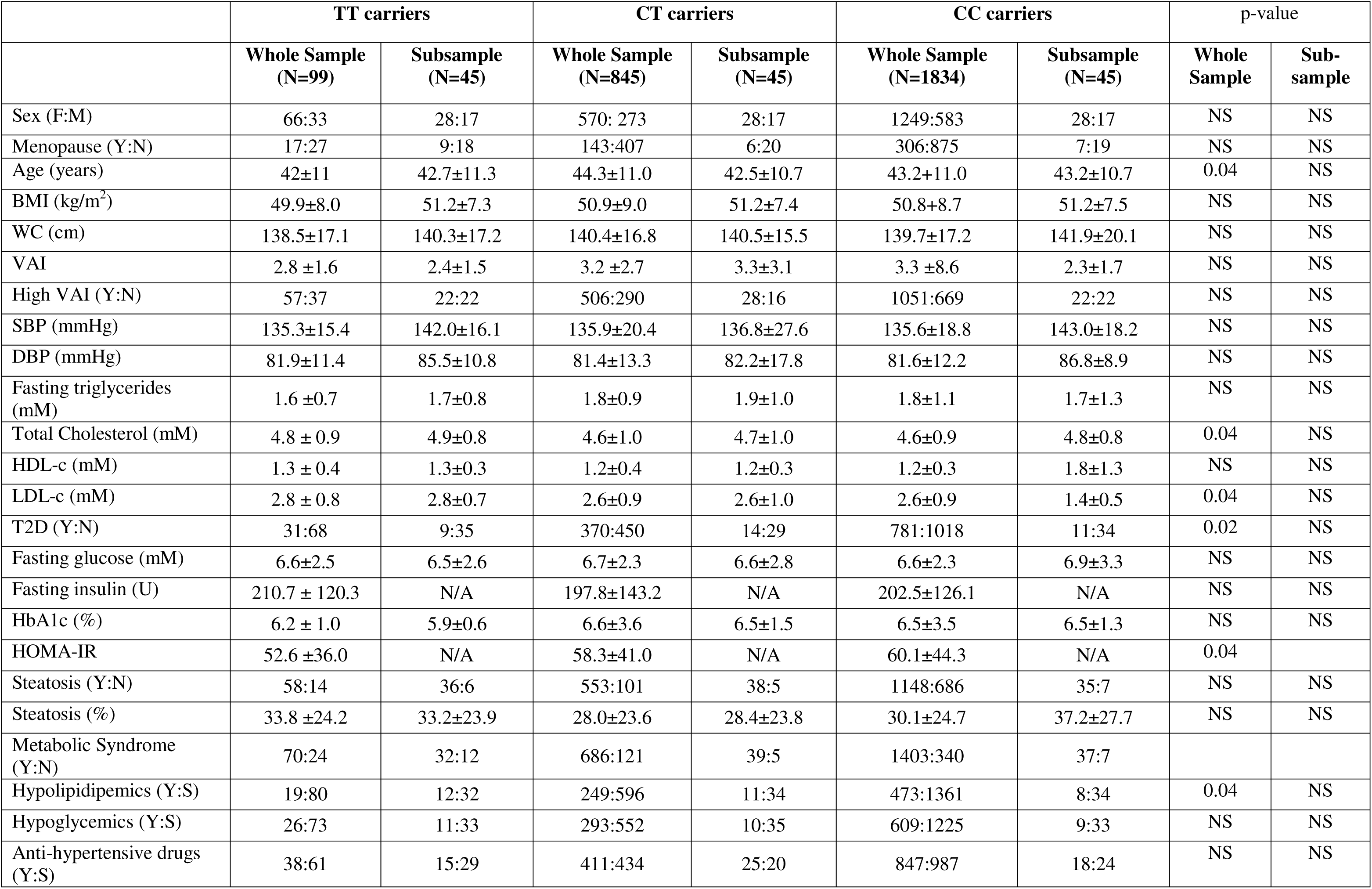
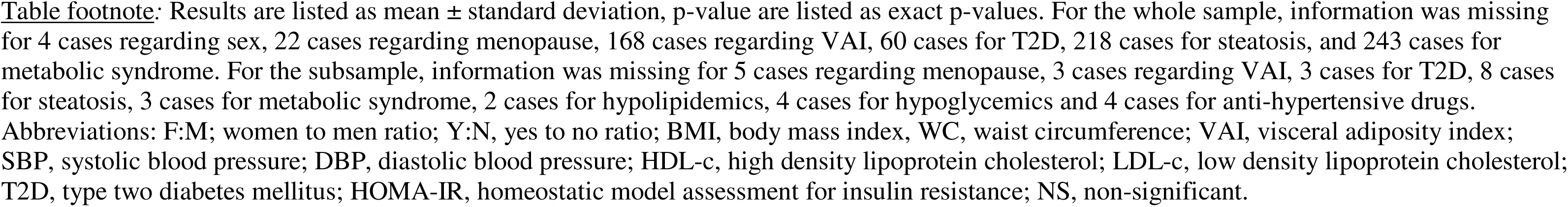
Characteristics of the study sample. Cardiometabolic risk profile of the whole sample (N=2798) as well as a subsample (N=135) of individuals with available adipose tissue and matched for age, sex, BMI and date of surgery.

We calculated the visceral adiposity index (22) for each participant using the following equation:

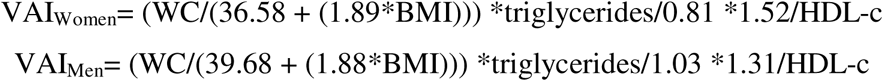

This study has been approved by the *Institut universitaire de cardiologie et pneumologie de Québec* (IUCPQ-Université Laval) Ethics Committee, according to the institutionally approved management modalities of its obesity biobank.

### Adipose tissue morphology

Subcutaneous and visceral adipose tissues samples were collected during surgery and flash frozen in liquid nitrogen directly in the operating room. Frozen samples were fixed for 24 hours at 4°C in 10% buffered formalin and then included in paraffin (23).

Following hematoxylin and eosin histological staining, slides were scanned using Axio Scan.Z1 Slide Scanner (ZEISS, Jena, Germany). Mean adipocyte diameter was computed using an automated in-house script in ImageJ (Rasband, W.S., ImageJ, U. S. National Institutes of Health, Bethesda, Maryland, USA, https://imagej.nih.gov/ij/, 1997-2018). Briefly, images were binarized using an automatic threshold to maximize contrast. In order to control for tissues imperfections (i.e. adipocytes with ruptured membranes or shrunken tissue) minimum and maximum size thresholds were set. Finally, adipocytes were considered imperfect circles and therefore given a circularity acceptance varying from 0.63 to 1 (1 being a perfect circle and 0 not being a circle). This process allowed us to measure an average of 819 adipocytes per slide (minimum = 31, maximum = 3340). Slides with less than 200 adipocytes or with mean adipocyte size more than two standard deviation above or below the mean were flagged for visual inspection.

We performed Picro-Sirius red staining on adipose tissue slides to assess pericellular fibrosis (24). Slides were scanned using Axio Scan.Z1 Slide Scanner (ZEISS, Jena, Germany) and analyzed using ImageJ using three internal standards to determine the batch-specific threshold (12).

### Gene expression

Whole, frozen adipose tissue (∼100 mg) was homogenized in QIAzol (Qiagen, Hilden, Germany) using Tissue Lyser (Qiagen, Hilden, Germany). RNeasy Lipid Tissue Mini Kit (Qiagen, Hilden, Germany) was used to extract RNA from adipocytes, which was then purified on columns through DNase digestion (Qiagen, Hilden, Germany). RNA was quantified using Biodrop (BioDrop Ltd., Cambridge, UK).

cDNA was then synthetized by the IScript reverse transcriptase (Bio-Rad Laboratories, Hercules, CA, USA), and real-time quantitative polymerase chain reaction (RT-qPCR) was conducted using SSO advance SYBR Green supermix transcriptase (Bio-Rad Laboratories, Hercules, CA, USA). An internal standard was used for calibration and signal threshold was manually set at the same level for both RT-qPCR plates measuring the expression of the same gene. NCBI primer design tool was used to design primers sequences (https://www.ncbi.nlm.nih.gov/tools/primer-blast/), while synthesis was performed by IDT (Integrated DNA Technology, Coralville, IA, USA). Primer’s specificity and annealing temperature were validated by gel electrophoresis in addition to melt curve analyses. The list of primers, including targets, sequences and annealing temperature are available in the supplementary data (**Table S1**).

### AKR1C activity

We measured intra-adipose AKR1C activity by fluorescence emission of coumberol, the reduced form of coumberone (25), as previously reported (12). Briefly, following tissue homogenization in 6.67 μL/mg potassium-phosphate buffer, we isolated the aqueous phase by centrifugation (20 624g, 16°C, 5 minutes). Reagents (10 nM NADPH, 25μM Coumberone) were then added to a 1:11 dilution. Fluorescence emission was monitored during 12-hour experiments (12). Blood and NADPH, which may interfere with the reading, were controlled for using a sample-specific blank. Enzymatic activity was calculated in the linear phase, i.e. using the slope of the first 30% of the curve (initial phase of the reaction).

### Statistical analyses

Statistical analyses were performed using R (Version 2023.06.0+421) (26). Assumptions were checked using *gvlma* package. When required, variables were transformed using the box-cox formula to reach normality. Planned-contrast ANOVA was carried on using *stats/ lme4* packages. The same packages were used to compute linear regressions, while Pearson’s correlations were done using the *car* package. In addition, mediation analyses were performed using *mediation* package. If not otherwise mentioned, data are represented as mean ± standard deviation (SD). Outliers were considered as values 2 standard deviation above the mean and were excluded from statistical analyses. Finally, to test the mode of inheritance, we tested the fit of linear regression in our models using the *lme4* package.

## RESULTS

According to public databases, rs28571848 has a MAF ranging between 6.4% (Qatari) and 22.5% (Northern Sweden). In our cohort, the MAF was at 18.7% close to the ones reported by TOPMED and TwinsUK (18.5% in both). In addition, rs28571848 has been suggested to increase *AKR1C2* and *AKR1C3* expression in both SAT and VAT adipose tissues (19). Therefore, one of our first objectives was to look at the expression of both enzymes in adipose tissue. As showed in **Figure 1**, rs28571848 significantly increased AKR1C3 expression in both SAT and VAT of women. This effect was not seen in men (**Figure 1A** and **1B**), which seemed to express an opposite phenotype (fold increase in “TT” homozygotes vs. “CC” homozygotes = 0.7, p-value >0.05). The SNP did not affect *AKR1C2* expression in adipose tissue (**Table 2**, **Figure 1C** and **1D**) (“TT” homozygotes vs. “CC” homozygotes fold increase in men =1.3 and in women=1, p-values >0.05). We additionally explored AKR1C activity. **Figure 1F** illustrates that women carrying the mutated “T” allele (i.e. “TT” homozygotes and “CT” heterozygotes) have increased AKR1C activity in VAT adipose tissue compared to “CC” homozygotes. We then tested whether the effects of the SNP followed a specific mode of inheritance. Our results showed that women *AKR1C3* expression in both adipose tissue depots follow the additive mode of inheritance model (for SAT: p-value_CC vs. TT_= 0.004; p-v; p-value_CT vs. TT_= 0.01; p-value_CC vs. CT_= NS; for VAT: p-value_CC vs. TT_= 0.03; p-v; p-value_CT vs. TT_= 0.01; p-value_CC vs. CT_= NS), while VAT AKR1C activity in women was dominant.

**Figure 1:**
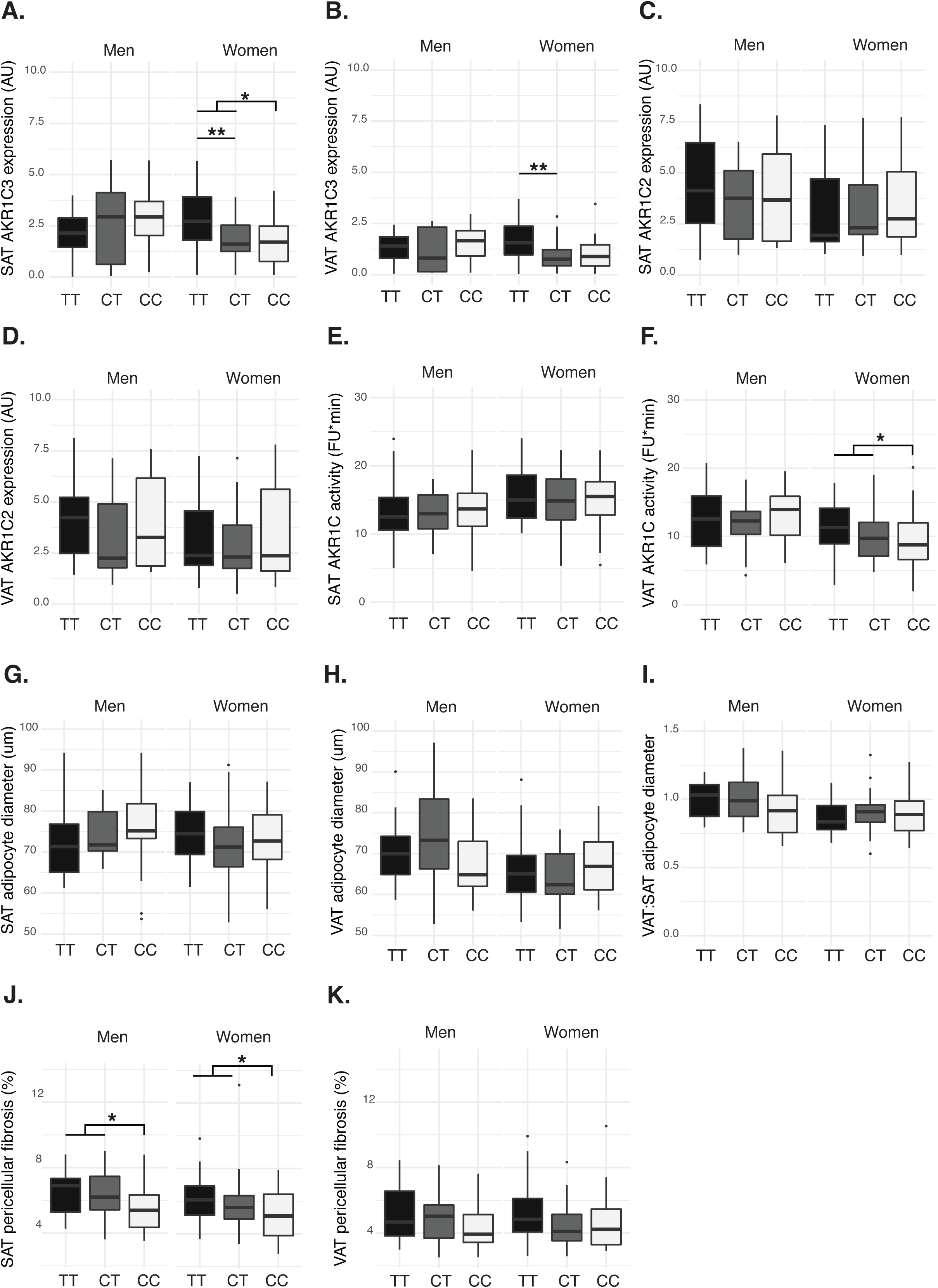
Adipose tissue features of the subsample of 135 individuals from which subcutaneous (SAT) and visceral adipose tissue (VAT) was obtained. Planned-contrast ANOVA was conducted in a sex-specific manner and significant differences (p-value <0.05) are reported. Abbreviations: TT, homozygous for the least frequent allele (T); CT, heterozygous; CC, homozygous for the most frequent allele (C). Gene expression is expressed in arbitrary units (AU) over the geometrical mean expression of two housekeeping genes (*ATP5O* and *HRPT1*). AKR1C activity is reported as fluorescent units (FU) multiplied by time, adipocyte mean diameter in micrometers (um) and pericellular fibrosis in percent.

**Table 2:**
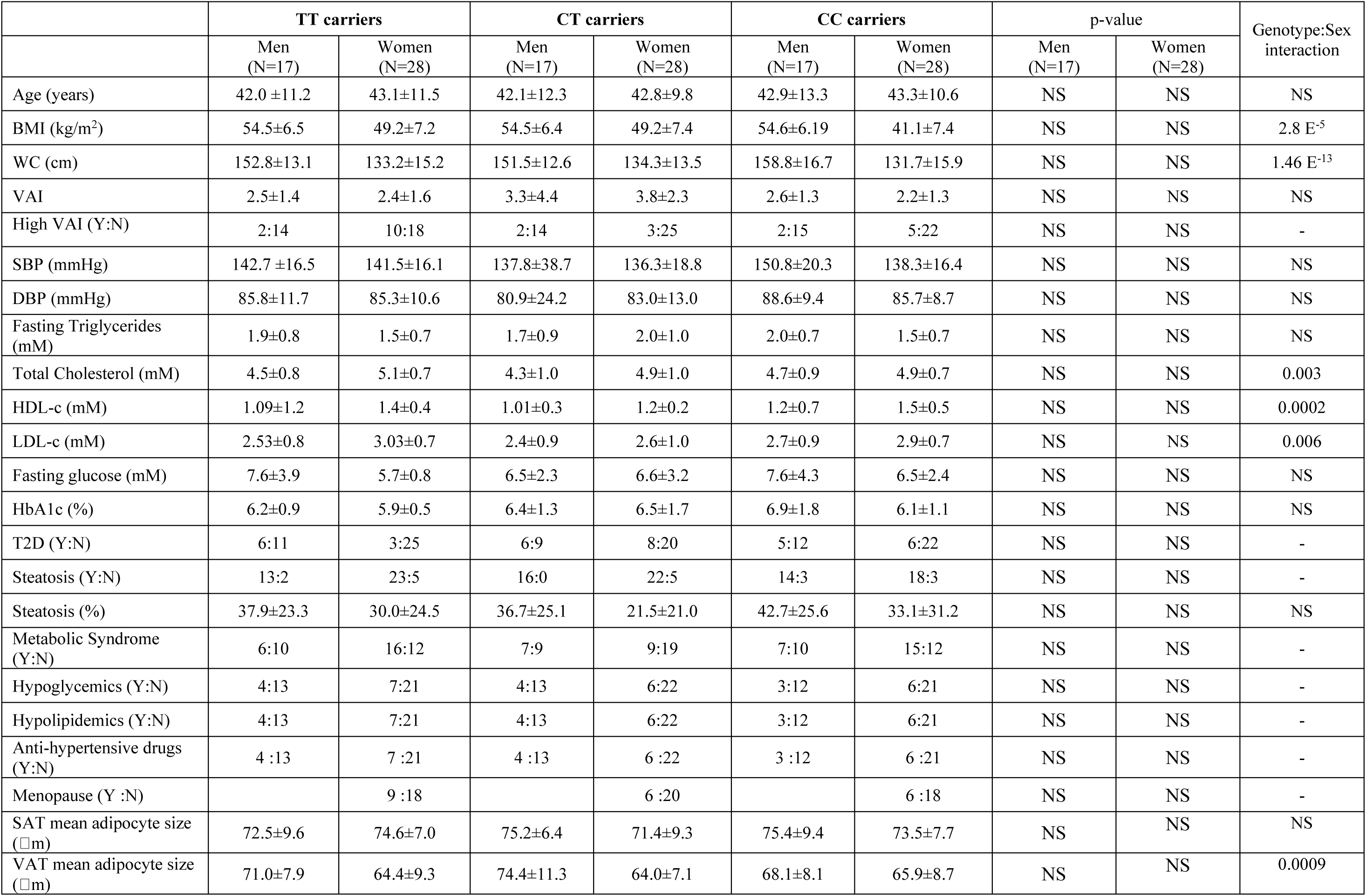
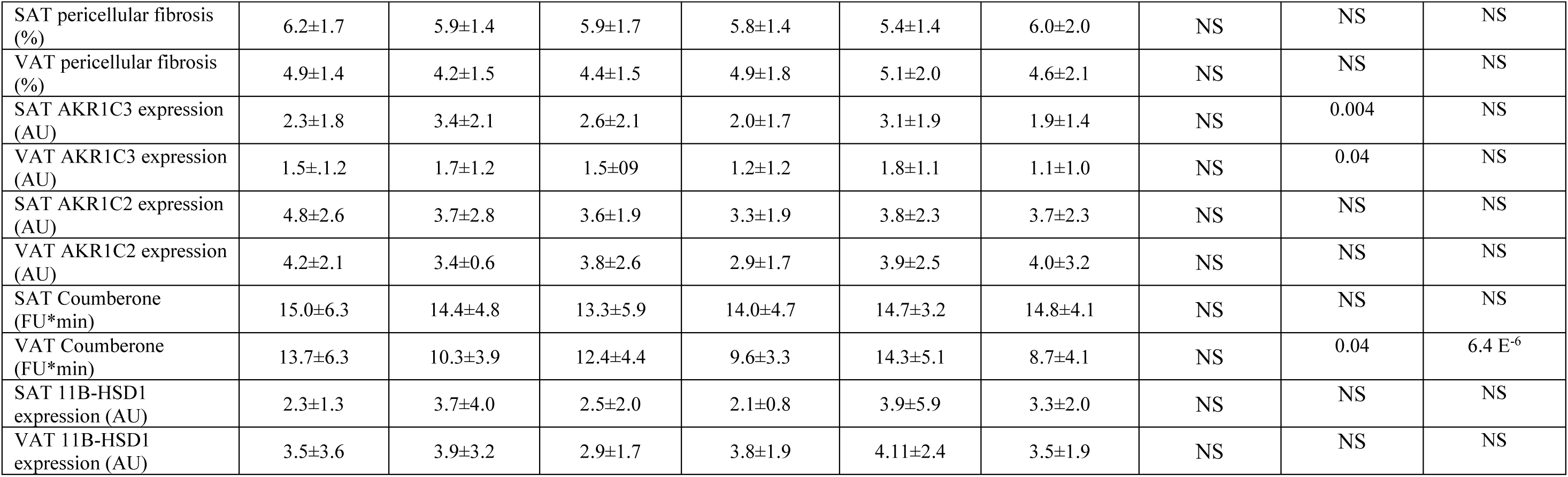
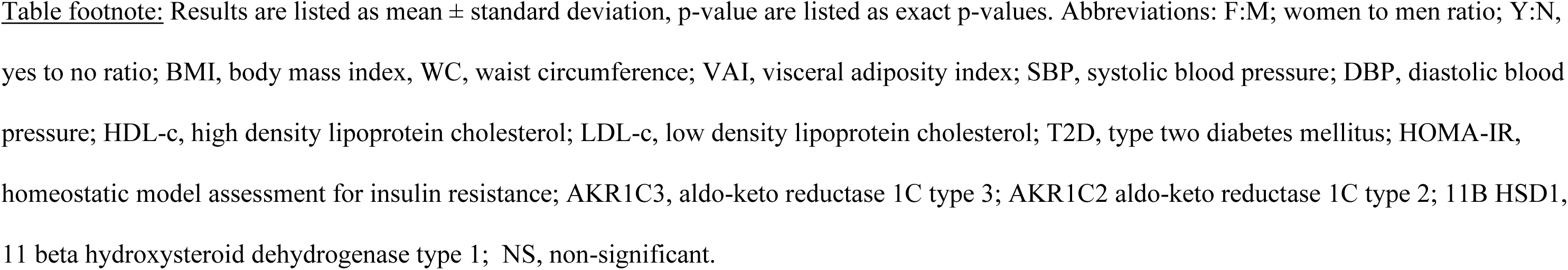
Metabolic and adipose tissue features of the subsample of 135 individuals. Comparison of the cardiometabolic profile, adipose tissue dysfunction features and expression or activity of the AKR1C between the three genotypes.

When comparing the cardiometabolic risk profile between the three groups, we found that “TT” carriers were characterized by an altered lipid profile compared to “CT” heterozygotes, showing raised total plasma cholesterol (**Figure 2B**) and LDL-c (**Figure 2D**), but not plasma triglycerides (**Figure 2A**) and HDL-c (**Figure 2C**). Total cholesterol and LDL-c persisted after adjustment for age, sex and BMI. We additionally analyzed glucose homeostasis in individuals without T2D and not taking any medication for glycemic control. Our results showed that although “T” allele carriers did not seem to show any difference in fasting glucose, HbA1c or insulin compared to “C” carriers, they displayed increased HOMA-IR (“CC” homozygous: 38.82 ± 19.58; “CT” heterozygous: 42.23 ± 21.83; “TT” homozygous: 49.87 ± 26.65) (**Figure 2E**), even after adjustment for age, sex and BMI. When stratified by sex, our analyses indicated that only women were affected by these cardiometabolic impairments (**Table 1**). Overall, the effect of the SNP on the metabolic traits seemed to follow the dominant mode of inheritance, rather than the additive mode of inheritance. Indeed, only LDL-c followed the additive model (p-value_CC vs. TT_= 0.04; p-v; p-value_CT vs. TT_= 0.02; p-value_CC vs. CT_= NS). To confirm the altered cardiometabolic profile in “T” allele carriers we investigated the prevalence of T2D, liver steatosis, and high visceral adiposity index [a marker of increased visceral adiposity (22, 27, 28)] according to sex (**Table 1**). We found no significant difference in VAI or in the prevalence of either T2D and liver steatosis.

**Figure 2:**
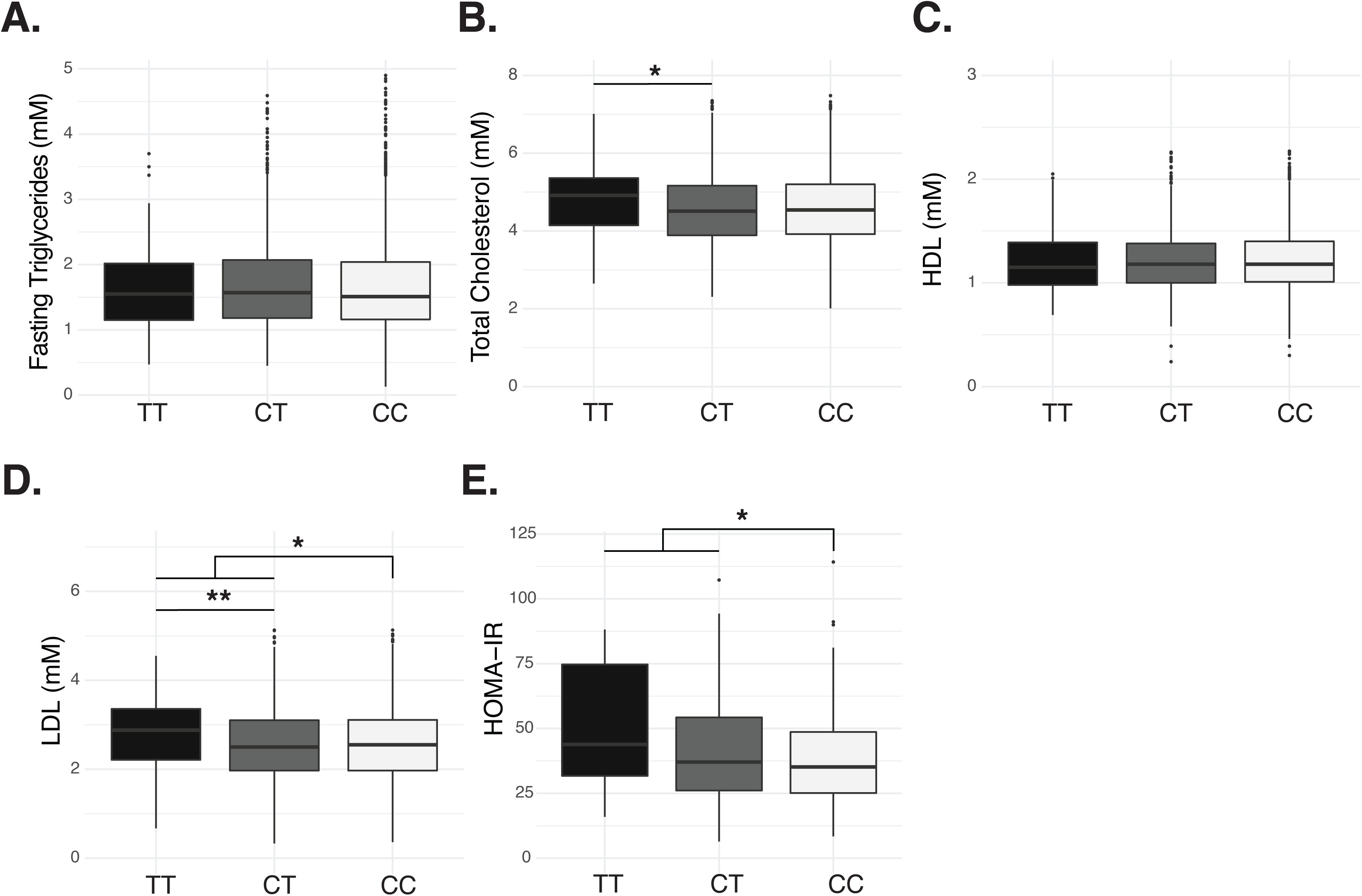
Sex-combined analysis of the cardiometabolic risk profile markers of the whole study sample (N=2798). Abbreviations: TT, homozygous for the least frequent allele (T); CT, heterozygous; CC, homozygous for the most frequent allele (C); HDL, high density lipoprotein cholesterol; LDL, low density lipoprotein cholesterol; HOMA-IR; homeostatic model assessment for insulin resistance.

We conducted correlation analyses between variables associated with cardiometabolic health, adipose tissue dysfunction, *AKR1C2* and *AKR1C3* gene expression and activity in adipose tissues of a subsample of 135 individuals where genotype groups were matched for BMI, age and sex (**Figure 3**). Our results showed a positive correlation between SAT mean adipocyte diameter and SAT *AKR1C3* expression (r=0.24, p=0.01). This association was confirmed by linear regression (β=1.08, p=0.01), which remained significant even after adjustment for sex (β=1.03, p<0.05). AKR1C activity in VAT was also positively correlated with a number of metabolic variables such as WC (r=0.21, p<0.05), HbA1c (r=0.28, p<0.05), fasting plasma triglycerides (r=0.20, p<0.05) and HDL-c (r=-0.27, p<0.01); as well as indices of adipose tissue dysfunction including VAT mean adipocyte diameter (r=0.23, p<0.01) and VAT pericellular fibrosis (r=0.22, p<0.05). While the association between WC and VAT adipocyte diameter was lost after adjustment for sex, there were trends for VAT fibrosis (p=0.06), plasma triglycerides (p=0.06), HDL-c (p=0.08) and HbA1c (p=0.06).

**Figure 3:**
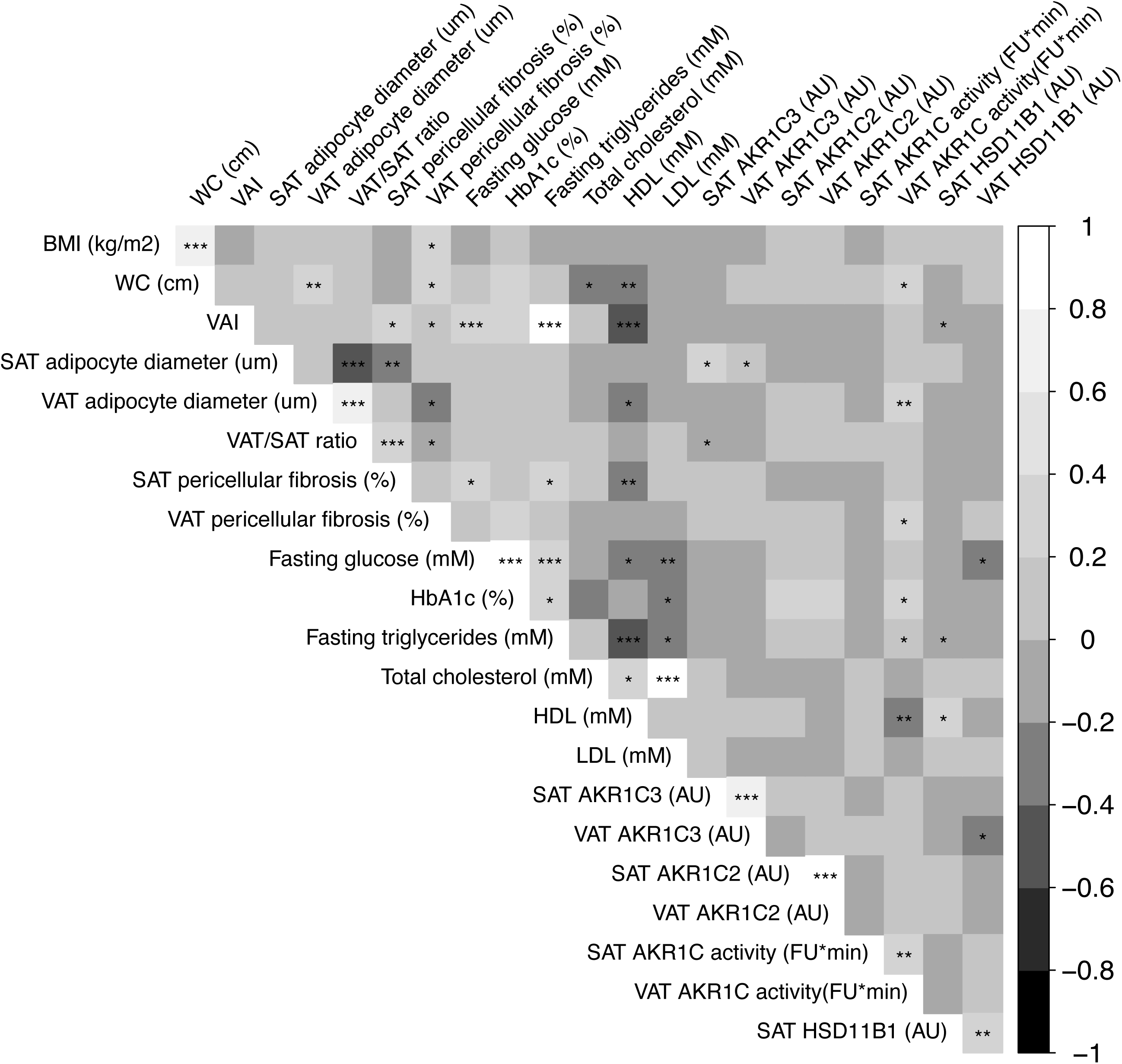
Pearson’s correlation plot of the subsample of 135 individuals from which subcutaneous (SAT) and visceral adipose tissue (VAT) was obtained. grey scale indicate the direction and strength of the correlation, while p-value represented by stars (*, p<0.05; **, p<0.01; ***, p<0.001). Abbreviations: WC, waist circumference; VAI, visceral adiposity index; SAT, subcutaneous adipose tissue; VAT, visceral adipose tissue; VAT/SAT ratio, VAT mean adipocyte diameter over SAT mean adipocyte diameter; HDL, high density lipoprotein cholesterol; LDL, low density lipoprotein cholesterol; AKR1C3, aldo keto reductase 1C3; AKR1C2; aldo keto reductase 1C2; HSD11B1, 11 beta hydroxysteroid dehydrogenase type 1.

Finally, to acquire additional insight on the impact that AKR1C might have on adipose tissue and cardiometabolic risk markers, we conducted mediation analyses and found that the effect seen of AKR1C VAT enzymatic activity on fasting triglycerides was partially mediated by VAT mean adipocyte size (mediation effect: β=0.009, p<0.05, proportion mediated = 18%).

## DISCUSSION

The objective of this study was to characterize the impact of AKR1C2 and AKR1C3 SNP rs28571848 on adipose tissue. Despite previous data identifying the “T” allele as a variant that increases the expression of both *AKR1C2* and *AKR1C3* in adipose tissue (19), our results show that only *AKR1C3* is affected. The observed increase in expression was corroborated by an increase in AKR1C activity, but in VAT only. Thus, rs28571848 is more likely to affect AKR1C3 expression and activity in adipose tissue rather than AKR1C2, and this particularly true in women. Hence, previous findings (19), based on open-source data (The Genotype-Tissue Expression project [https://www.gtexportal.org/home/]), were not confirmed by this study.

In addition, we found a MAF of 18.7%, which is closer to the one reported by TwinsUK than that of the 1000 genome project (19). This might be due to the fact that the 1000 Genome Project (29) is composed of approximately the same sample size as the one used in this study (i.e. 2504 vs. 2776 individuals) but issued from 26 different populations, while here, the participants were recruited were from a more homogeneous population. Because SNP frequency may vary between populations (30, 31), the MAF of rs28571848 in Caucasians might have been slightly underestimated by the 1000 Genome Project.

“TT” homozygotes additionally displayed altered cardiometabolic health with increased plasma LDL-cholesterol, total cholesterol, and HOMA-IR. Further confirming the association with cardiometabolic health, our correlation analyses revealed a positive correlation between AKR1C activity in VAT and WC, HbA1c and plasma triglycerides and a negative association with plasma HDL-c. Interestingly, features of adipose tissue dysfunction such as adipocyte mean diameter and pericellular fibrosis were associated with AKR1C3 expression or AKR1C activity in adipose tissue.

In the literature, AKR1C2 and AKR1C3 have been associated to central fat distribution (32), and this association is still reported by more recent studies using cutting-edge technologies. Using genome-wide transcriptional technologies, we found that *AKR1C2* and *AKR1C3* gene expression in SAT was positively correlated with trunk fat mass (19). Similarly, a study conducted on BMI-discordant twin women found that *AKR1C2* and *AKR1C3* gene expression was increased in the twin with a higher BMI (17, 18). The same group additionally found that within male twin pairs (i.e. independently of genetics), intra-abdominal adipose tissue as well as the intra-abdominal to subcutaneous fat-ratio were positively associated with the expression of *AKR1C2* and *AKR1C3* (18). Increased visceral adiposity has been associated with increased cardiometabolic risk (33) and the amount of VAT adjusted for BMI is consistently associated with increased T2D and cardiovascular risk in both sexes and across BMI categories (34). Although causality cannot be established, these data suggest that the adipose tissue hormonal milieu may be tightly linked to visceral adiposity and cardiometabolic health.

In adipose tissue, while AKR1C2 is known to be involved in the catabolism of the potent androgen DHT, AKR1C3 catalyzes the reaction between androstenedione and testosterone (35, 36), but can also yield small concentrations of estradiol (36) and has been implicated in the conversion of keto androgens (37, 38). Increased testosterone resulting from increased *AKR1C3* gene expression in adipocytes is known to be intrinsically linked to increased lipid accumulation in human adipose tissue. Indeed, Svensson and collaborators (39) performed a microarray study using human adipocytes that were previously isolated and separated by their size. Their results suggested that large adipocytes had a 1.5-fold increase in the expression of *AKR1C3* compared to the smaller adipocyte counterpart from the same participant (39). In addition, studies conducted with *in vitro* differentiated human adipocytes suggest that incubation with either testosterone or DHT increases *de novo* lipogenesis but not fatty acid uptake (40). This was further confirmed by the increased acetyl-CoA carboxylase alpha expression observed after testosterone or DHT administration (40). Because acetyl-CoA carboxylase alpha is the rate limiting enzyme in the lipogenic cascade (41), its increase suggests that androgens, such as testosterone, might facilitate *de novo* lipogenesis in adipose tissue. Although we could not find any significant difference in mean adipocyte size between the three genotype groups, we found a trend for higher VAT:SAT mean adipocyte diameter ratio in men carrying the “T” allele and a significant positive correlation between VAT diameter and VAT AKR1C activity. Adipocyte cell size is known to increase across BMI categories, but this relationship reaches a plateau in the obese state (42). The mechanisms behind adipocyte hypertrophy and altered adipose function are numerous (43), and despite literature suggesting lower SAT adipogenic capacity (44, 45), genome-wide association studies (GWAS) were unsuccessful in identifying gene variations associated with adipocyte size (46). This lack of significant results is probably due to the high heterogeneity of the molecular mechanisms implicated in adipocyte hypertrophy. Therefore, in this multifactorial landscape where adipocyte diameter is likely to result from multiple factors and where mean adipocyte diameter has probably reached its plateau, the lack of significant difference between the three genotype groups is not surprising. In addition, this study is based on the evaluation of a single SNP found in a region of high linkage disequilibrium (19), other SNP may have stronger effects. Nevertheless, the presence of a positive association between SAT adipocyte diameter and SAT *AKR1C3* gene expression, still is significant, suggesting that the adipose tissue hormonal milieu plays an important role in adipose tissue function, independently of genotype.

Hypertrophic adipocytes, one of the major hallmark of adipose tissue dysfunction, are known to inefficiently store fat (2), significantly increasing lipotoxicity in the lean organs. Adipocyte *AKR1C3* gene expression has already been linked to lipotoxicity in humans (40, 47). In the present study, not only did we find AKR1C3 expression and activity to be associated with mean adipocyte diameter in VAT, but our mediation model also suggested that the effects of VAT AKR1C activity on fasting plasma triglycerides were partially mediated by adipocyte size. Therefore, we suggest that rs28571848 increases AKR1C3 expression and activity in adipose tissue, facilitating lipid accumulation and adipocyte hypertrophy. The establishment of adipose tissue dysfunction will subsequently lead to impaired cardiometabolic health.

The present study has a number of limitations and therefore it should be interpreted with caution. First, in-depth characterization of adipose tissue was conducted only on 135 participants (84 women, 51 men). This was due to the availability of adipose tissue (especially in “TT” homozygotes) and our selective matching criteria. The limited sample size may have affected power, especially as far as sex-specific analyses are concerned. Here, we showed that rs28571848 affected *AKR1C3* gene expression in women, but not in men. We cannot exclude the possibility that with a larger sample size, differences in *AKR1C3* gene expression would be confirmed in men. In addition, we were unable to show differences in *AKR1C2* gene expression. Because the effects on *AKR1C2* gene expression might be of small magnitude, a much greater sample size might be required to contrast possible factors affecting variance. These limitations are due to the fact that adipose tissue characterization was conducted on a limited sample. However, we were able to retrieve medical records of a large number of patients. This allowed us to identify differences in plasma cholesterol and insulin resistance and therefore further characterize the metabolic impairments found in connection with rs28571848.

Second, as mentioned before, our study sample is composed of bariatric surgery candidates with severe obesity and obesity-associated comorbidities. Although we controlled for this by conducting analyses on glucose homeostasis excluding individuals taking hypoglycemics or with diagnosed T2D, a possible confounding effect cannot be excluded. In addition, adipocyte size of individuals with severe obesity is likely to have reached a plateau (42), and therefore the effects of a single factor may be much more difficult to isolate. The inclusion of individuals without obesity might have revealed stronger effects of rs28571848. In this study, an attempt to control for the confounding effect of cardiometabolic health in severe obesity was made by including a control group (“CC” homozygotes) issued from the same population and therefore having comparable characteristics as the cases (“TT” homozygotes).

Finally, to measure AKR1C activity in adipose tissue we used coumberone, a molecule that when converted into coumberol becomes fluorescent (25). Although originally coumberone was developed as a specific substrate of AKR1C2 (25), later studies suggested that it displays affinities for all the AKR1C family members (48, 49). AKR1C family shares high similarity both in terms of genetic sequence and amino acids identity, but strong differences for substrate preferences (14, 35, 47). As far as adipose tissue is concerned, this means that coumberone measurements assess AKR1C1, AKR1C2 and AKR1C3 activities but not AKR1C4, as this enzyme is mainly expressed in the liver and the gall bladder (15). Supporting this notion, previous studies identified *AKR1C2* as the most highly expressed enzyme of the AKR1C family in adipose tissue (32), followed by *AKR1C3* and *AKR1C1*, while *AKR1C4* is not detected (32). AKR1C1 is involved in progesterone catabolism (47) and has a relatively low affinity for androgens (35, 47), while AKR1C3 and AKR1C2 are involved in testosterone synthesis and DHT degradation respectively (14, 35, 47). Previous studies suggested that AKR1C3 is highly labile, and upon homogenization its detectable activity drops by 90% reaching approximately 10% of that measurable in intact cells (50). However, our results seem to follow the expression of *AKR1C3* rather than *AKR1C2* suggesting that AKR1C3 activity might have been detected despite homogenization.

In conclusion, we found that rs28571858 affects adipose tissue *AKR1C3* expression in women and that its expression is mirrored by an increased AKR1C activity. Increased AKR1C3 may participate in the creation of an adipose tissue hormonal milieu that favors adipose tissue expansion and alters cardiometabolic health by deteriorating lipid homeostasis and insulin sensitivity.

## Data Availability

The participants of this study did not give written consent for their data to be shared publicly, so due to the sensitive nature of the research, raw data are not available.

## ACKNOWLEDGMENTS

The authors recognize the help of the CRIUCPQ Biobank as well as all the surgical team assisting bariatric surgery patients during the procedure and collection of adipose tissues samples. GO’s current affiliation is at Montreal University Hospital Research Center (CRCHUM), Montréal (QC), Canada affiliated to Montreal University.

## REFERENCES

1. Tchernof A, Després JP. Pathophysiology of human visceral obesity: an update. Physiol Rev. 2013;93(1):359–404.

2. Neeland IJ, Ross R, Després JP, Matsuzawa Y, Yamashita S, Shai I, et al. Visceral and ectopic fat, atherosclerosis, and cardiometabolic disease: a position statement. Lancet Diabetes Endocrinol. 2019;7(9):715–25.

3. Rao VN, Bush CG, Mongraw-Chaffin M, Hall ME, Clark D, 3rd, Fudim M, et al. Regional Adiposity and Risk of Heart Failure and Mortality: The Jackson Heart Study. J Am Heart Assoc. 2021;10(14):e020920.

4. Rosito GA, Massaro JM, Hoffmann U, Ruberg FL, Mahabadi AA, Vasan RS, et al. Pericardial fat, visceral abdominal fat, cardiovascular disease risk factors, and vascular calcification in a community-based sample: the Framingham Heart Study. Circulation. 2008;117(5):605–13.

5. Neeland IJ, Turer AT, Ayers CR, Powell-Wiley TM, Vega GL, Farzaneh-Far R, et al. Dysfunctional adiposity and the risk of prediabetes and type 2 diabetes in obese adults. Jama. 2012;308(11):1150–9.

6. Gagnon E, Mitchell PL, Arsenault BJ. Body fat distribution, fasting insulin levels and insulin secretion: A bidirectional Mendelian randomization study. J Clin Endocrinol Metab. 2022.

7. Gagnon E, Pelletier W, Gobeil É, Bourgault J, Manikpurage HD, Maltais-Payette I, et al. Mendelian randomization prioritizes abdominal adiposity as an independent causal factor for liver fat accumulation and cardiometabolic diseases. Commun Med (Lond). 2022;2:130.

8. Tchernof A, Brochu D, Maltais-Payette I, Mansour MF, Marchand GB, Carreau AM, et al. Androgens and the Regulation of Adiposity and Body Fat Distribution in Humans. Compr Physiol. 2018;8(4):1253–90.

9. Ruth KS, Day FR, Tyrrell J, Thompson DJ, Wood AR, Mahajan A, et al. Using human genetics to understand the disease impacts of testosterone in men and women. Nat Med. 2020;26(2):252–8.

10. Loh NY, Humphreys E, Karpe F, Tomlinson JW, Noordam R, Christodoulides C. Sex hormones, adiposity, and metabolic traits in men and women: a Mendelian randomisation study. Eur J Endocrinol. 2022;186(3):407–16.

11. Tchernof A, Mansour MF, Pelletier M, Boulet MM, Nadeau M, Luu-The V. Updated survey of the steroid-converting enzymes in human adipose tissues. J Steroid Biochem Mol Biol. 2015;147:56–69.

12. Ostinelli G, Laforest S, Denham SG, Gauthier MF, Drolet-Labelle V, Scott E, et al. Increased Adipose Tissue Indices of Androgen Catabolism and Aromatization in Women With Metabolic Dysfunction. J Clin Endocrinol Metab. 2022;107(8):e3330–e42.

13. Colldén H, Nilsson ME, Norlén AK, Landin A, Windahl SH, Wu J, et al. Comprehensive Sex Steroid Profiling in Multiple Tissues Reveals Novel Insights in Sex Steroid Distribution in Male Mice. Endocrinology. 2022;163(3).

14. Dufort I, Labrie F, Luu-The V. Human types 1 and 3 3 alpha-hydroxysteroid dehydrogenases: differential lability and tissue distribution. J Clin Endocrinol Metab. 2001;86(2):841–6.

15. Fagerberg L, Hallström BM, Oksvold P, Kampf C, Djureinovic D, Odeberg J, et al. Analysis of the human tissue-specific expression by genome-wide integration of transcriptomics and antibody-based proteomics. Mol Cell Proteomics. 2014;13(2):397–406.

16. Blouin K, Nadeau M, Mailloux J, Daris M, Lebel S, Luu-The V, et al. Pathways of adipose tissue androgen metabolism in women: depot differences and modulation by adipogenesis. Am J Physiol Endocrinol Metab. 2009;296(2):E244–55.

17. Vihma V, Heinonen S, Naukkarinen J, Kaprio J, Rissanen A, Turpeinen U, et al. Increased body fat mass and androgen metabolism - A twin study in healthy young women. Steroids. 2018;140:24–31.

18. Vihma V, Naukkarinen J, Turpeinen U, Hämäläinen E, Kaprio J, Rissanen A, et al. Metabolism of sex steroids is influenced by acquired adiposity-A study of young adult male monozygotic twin pairs. J Steroid Biochem Mol Biol. 2017;172:98–105.

19. Ostinelli G, Vijay J, Vohl MC, Grundberg E, Tchernof A. AKR1C2 and AKR1C3 expression in adipose tissue: Association with body fat distribution and regulatory variants. Mol Cell Endocrinol. 2021;527:111220.

20. Blouin K, Blanchette S, Richard C, Dupont P, Luu-The V, Tchernof A. Expression and activity of steroid aldoketoreductases 1C in omental adipose tissue are positive correlates of adiposity in women. Am J Physiol Endocrinol Metab. 2005;288(2):E398–404.

21. Blouin K, Richard C, Bélanger C, Dupont P, Daris M, Laberge P, et al. Local androgen inactivation in abdominal visceral adipose tissue. J Clin Endocrinol Metab. 2003;88(12):5944–50.

22. Amato MC, Giordano C, Galia M, Criscimanna A, Vitabile S, Midiri M, et al. Visceral Adiposity Index: a reliable indicator of visceral fat function associated with cardiometabolic risk. Diabetes Care. 2010;33(4):920–2.

23. Laforest S, Pelletier M, Michaud A, Daris M, Descamps J, Soulet D, et al. Histomorphometric analyses of human adipose tissues using intact, flash-frozen samples. Histochem Cell Biol. 2018;149(3):209–18.

24. Michaud A, Tordjman J, Pelletier M, Liu Y, Laforest S, Noël S, et al. Relevance of omental pericellular adipose tissue collagen in the pathophysiology of human abdominal obesity and related cardiometabolic risk. Int J Obes (Lond). 2016;40(12):1823–31.

25. Yee DJ, Balsanek V, Bauman DR, Penning TM, Sames D. Fluorogenic metabolic probes for direct activity readout of redox enzymes: Selective measurement of human AKR1C2 in living cells. Proc Natl Acad Sci U S A. 2006;103(36):13304–9.

26. R Core Team. R: A Language and Environment for Statistical Computing. R Foundation for Statistical Computing, Vienna. 2013.

27. Amato MC, Giordano C. Visceral adiposity index: an indicator of adipose tissue dysfunction. Int J Endocrinol. 2014;2014:730827.

28. Amato MC, Giordano C, Pitrone M, Galluzzo A. Cut-off points of the visceral adiposity index (VAI) identifying a visceral adipose dysfunction associated with cardiometabolic risk in a Caucasian Sicilian population. Lipids Health Dis. 2011;10:183.

29. Fairley S, Lowy-Gallego E, Perry E, Flicek P. The International Genome Sample Resource (IGSR) collection of open human genomic variation resources. Nucleic Acids Res. 2020;48(D1):D941–d7.

30. Cheung EYY, Phillips C, Eduardoff M, Lareu MV, McNevin D. Performance of ancestry-informative SNP and microhaplotype markers. Forensic Sci Int Genet. 2019;43:102141.

31. Stoneking M, Krause J. Learning about human population history from ancient and modern genomes. Nat Rev Genet. 2011;12(9):603–14.

32. Wake DJ, Strand M, Rask E, Westerbacka J, Livingstone DE, Soderberg S, et al. Intra-adipose sex steroid metabolism and body fat distribution in idiopathic human obesity. Clin Endocrinol (Oxf). 2007;66(3):440–6.

33. Piché ME, Tchernof A, Després JP. Obesity Phenotypes, Diabetes, and Cardiovascular Diseases. Circ Res. 2020;126(11):1477–500.

34. Agrawal S, Klarqvist MDR, Diamant N, Stanley TL, Ellinor PT, Mehta NN, et al. BMI-adjusted adipose tissue volumes exhibit depot-specific and divergent associations with cardiometabolic diseases. Nat Commun. 2023;14(1):266.

35. Veilleux A, Côté JA, Blouin K, Nadeau M, Pelletier M, Marceau P, et al. Glucocorticoid-induced androgen inactivation by aldo-keto reductase 1C2 promotes adipogenesis in human preadipocytes. Am J Physiol Endocrinol Metab. 2012;302(8):E941–9.

36. Penning TM, Byrns MC. Steroid hormone transforming aldo-keto reductases and cancer. Ann N Y Acad Sci. 2009;1155:33–42.

37. Paulukinas RD, Mesaros CA, Penning TM. Conversion of Classical and 11-Oxygenated Androgens by Insulin-Induced AKR1C3 in a Model of Human PCOS Adipocytes. Endocrinology. 2022;163(7).

38. Barnard M, Quanson JL, Mostaghel E, Pretorius E, Snoep JL, Storbeck KH. 11-Oxygenated androgen precursors are the preferred substrates for aldo-keto reductase 1C3 (AKR1C3): Implications for castration resistant prostate cancer. J Steroid Biochem Mol Biol. 2018;183:192–201.

39. Svensson PA, Gabrielsson BG, Jernås M, Gummesson A, Sjöholm K. Regulation of human aldoketoreductase 1C3 (AKR1C3) gene expression in the adipose tissue. Cell Mol Biol Lett. 2008;13(4):599–613.

40. O’Reilly MW, Kempegowda P, Walsh M, Taylor AE, Manolopoulos KN, Allwood JW, et al. AKR1C3-Mediated Adipose Androgen Generation Drives Lipotoxicity in Women With Polycystic Ovary Syndrome. J Clin Endocrinol Metab. 2017;102(9):3327–39.

41. Munday MR. Regulation of mammalian acetyl-CoA carboxylase. Biochem Soc Trans. 2002;30(Pt 6):1059–64.

42. Laforest S, Labrecque J, Michaud A, Cianflone K, Tchernof A. Adipocyte size as a determinant of metabolic disease and adipose tissue dysfunction. Crit Rev Clin Lab Sci. 2015;52(6):301–13.

43. Stenkula KG, Erlanson-Albertsson C. Adipose cell size: importance in health and disease. Am J Physiol Regul Integr Comp Physiol. 2018;315(2):R284–r95.

44. Lessard J, Laforest S, Pelletier M, Leboeuf M, Blackburn L, Tchernof A. Low abdominal subcutaneous preadipocyte adipogenesis is associated with visceral obesity, visceral adipocyte hypertrophy, and a dysmetabolic state. Adipocyte. 2014;3(3):197–205.

45. Ledoux S, Boulet N, Belles C, Zakaroff-Girard A, Bernard A, Germain A, et al. Subcutaneous Stromal Cells and Visceral Adipocyte Size Are Determinants of Metabolic Flexibility in Obesity and in Response to Weight Loss Surgery. Cells. 2022;11(22).

46. Glastonbury CA, Pulit SL, Honecker J, Censin JC, Laber S, Yaghootkar H, et al. Machine Learning based histology phenotyping to investigate the epidemiologic and genetic basis of adipocyte morphology and cardiometabolic traits. PLoS Comput Biol. 2020;16(8):e1008044.

47. Penning TM, Wangtrakuldee P, Auchus RJ. Structural and Functional Biology of Aldo-Keto Reductase Steroid-Transforming Enzymes. Endocr Rev. 2019;40(2):447–75.

48. Guise CP, Abbattista MR, Singleton RS, Holford SD, Connolly J, Dachs GU, et al. The bioreductive prodrug PR-104A is activated under aerobic conditions by human aldo-keto reductase 1C3. Cancer Res. 2010;70(4):1573–84.

49. Jamieson SM, Gu Y, Manesh DM, El-Hoss J, Jing D, Mackenzie KL, et al. A novel fluorometric assay for aldo-keto reductase 1C3 predicts metabolic activation of the nitrogen mustard prodrug PR-104A in human leukaemia cells. Biochem Pharmacol. 2014;88(1):36–45.

50. Dufort I, Rheault P, Huang XF, Soucy P, Luu-The V. Characteristics of a highly labile human type 5 17beta-hydroxysteroid dehydrogenase. Endocrinology. 1999;140(2):568–74.

