## Supplemental Table 1 for "Impact of *AKR1C2* and *AKR1C3* single nucleotide polymorphism rs28571848 in adipose tissues of individuals with severe obesity"

**Table S1: List of primers used**. Primers were designed using NCBI primer design tool and validated by PCR before use.

| **Gene** | **GenBank reference** | **Size (bp)** | **Sequence** (5‘ → 3‘// 3‘→5‘) |
|---|---|---|---|
| ATP5O | NM_001697 | 267 | ATTGAAGGTCGCTATGCCACAG// AACGACTCCTTGGGTATTGCTTAA |
| HMBS | NM_00190.4 | 235 | TGCAGAGAAAGTTCCCGCAT// AAGATGTCCTGGTCCTTGGC |
| AKR1C2 | NM_001354.6 | 341 | GCTCTTATAGCCTGTGAGGGAG // GACCAACTCTGGTCGATGGG |
| AKR1C3 | NM_003739.6 | 150 | ACAGGGAATGGATTCCAAACACCA // TGGCGGAACCCAGCTTCTAT |
| HSD11B1 | NM_005525.4 | 269 | CATTGCTGGCACCATGGAAG// GATAAGCCACTTTCCCAGCCA |
